# Patients with presumed tuberculosis in sub-Saharan Africa that are not diagnosed with tuberculosis: a systematic review and meta-analysis

**DOI:** 10.1101/2021.05.19.21257444

**Authors:** S Jayasooriya, F Dimambro-Denson, C Beecroft, J Balen, B Awokola, C Mitchell, B Kampmann, F Campbell, PJ Dodd, K Mortimer

## Abstract

**Background:** Many patients in sub-Saharan Africa whom a diagnosis of tuberculosis is considered are subsequently not diagnosed with tuberculosis. The proportion of patients this represents, and their alternative diagnoses, have not previously been systematically reviewed.

**Methods:** We searched four databases from inception to April 27, 2020, without language restrictions (PROSPERO: CRD42018100004). We included all adult pulmonary tuberculosis diagnostic studies from sub-Saharan Africa, excluding case series and inpatient studies. We extracted the proportion of patients with presumed tuberculosis subsequently not diagnosed with tuberculosis and any alternative diagnoses received. We conducted a random-effects meta-analysis to obtain pooled estimates stratified by passive and active case finding.

**Results:** Our search identified 1799 studies, of which 18 studies with 14527 participants from 10 African countries were included. The proportion of patients with presumed tuberculosis subsequently not diagnosed with tuberculosis was 48.5% (95% CI 38.4-56.7) in passive and 92.7% (95% CI 83.1-97.0) in active case finding studies. This proportion increased with declining numbers of clinically diagnosed tuberculosis cases. Past history of tuberculosis was documented in only 55% of studies, with just five out of 18 reporting any alternative diagnoses.

**Discussion:** Nearly half of all patients with presumed tuberculosis in sub-Saharan Africa do not have a final diagnosis of active tuberculosis. This proportion may be higher when active case finding strategies are used. Little is known about the healthcare needs of these patients. Research is required to better characterise these patient populations and plan health system solutions that meet their needs.

**Funding:** NIHR, UK MRC

## Introduction

The differential access to high quality diagnostics experienced in most low- middle-income countries (LMICs) illustrate important and growing global health disparities.^1^ Diagnostic tests are often not accessible, affordable or designed for application in LMICs forming a barrier to providing high-quality healthcare. Access to accurate diagnostics for a range of diseases is a cornerstone of high-quality patient care, enabling appropriate timely management, inclusive of transmission control in the case of communicable disease. Pulmonary tuberculosis is a highly prevalent poverty-related communicable disease that lays bare many of the diagnostic challenges faced in LMICs, not least because of non-specific symptoms at presentation.^2^

Patients with presumed tuberculosis are adults or children evaluated for active tuberculosis with suggestive signs and symptoms, such as cough, fever, night sweats, weight loss, haemoptysis and fatigue. While sputum culture remains the bacteriological reference standard for tuberculosis diagnostics, it is a costly, lengthy process and in LMICs is usually only available in central reference laboratories. At local clinics, a reliance on smear microscopy is being replaced by rapid diagnostics such as Xpert MTB/RIF and Xpert MTB/RIF Ultra nucleic acid amplification tests (sensitivity 85%, specificity 98%).^3^ Despite these advances, only 57% of global pulmonary tuberculosis cases are bacteriologically confirmed, the rest are clinically diagnosed with negative or no bacteriological testing and notified to the World Health Organisation (WHO) as such. Whereas in high income settings 80% of pulmonary tuberculosis cases are confirmed bacteriologically.^4^ The WHO describes the use of both passive and active case finding strategies to detect tuberculosis cases.^2^ Passive case finding relies on symptomatic patients seeking medical care by presenting to health services, whereas active case finding involves community-based screening of high risk patients who would not otherwise seek healthcare.

A proportion of patients with presumed tuberculosis are found not to have tuberculosis, following both bacteriological and clinical investigation. This proportion is likely to depend on tuberculosis prevalence, case finding strategies (passive or active) and other context-specific factors such as access to alternative diagnostics. A community study in Malawi demonstrated that only 10-20% of patients presenting to primary care with a persistent cough had tuberculosis.^5^ More recent observational data from The Gambia^6^ showed that nearly half of all patients with presumed tuberculosis did receive a final diagnosis of tuberculosis. A range of alternative diagnoses - predominantly respiratory - were described, but importantly, non-respiratory diagnoses such as heart failure, malignancy and renal failure were also noted. Furthermore, in 36% of patients not diagnosed with tuberculosis, no alternative diagnosis was made. Minimal health care was afforded to these patients beyond screening for tuberculosis and Human Immunodeficiency Virus (HIV). The burden of ill-health in patients with presumed tuberculosis subsequently found not to have tuberculosis and their on-going engagement with health systems has been largely overlooked. While national guidelines exist for patients that receive a negative sputum smear microscopy result, these focus on further elucidating active tuberculosis cases rather than exploring alternative diagnoses. The rapid rise of non-communicable disease – including chronic respiratory diseases^1^ – in tuberculosis endemic areas, means patients presenting with presumed tuberculosis may increasingly have alternative health issues that require investigation and management, once tuberculosis is ruled out.

Since the Alma Ata declaration, recognising primary healthcare as key to attaining the goal of ‘health for all’, there have been several strategies addressing lung health through service integration and guidance. These include the Practical Approach to Lung Health (PAL) WHO strategy,^7^ and The Practical Approach to Care Kit (PACK).^8^ The former advocates for the integration of alternative diagnostics for other respiratory conditions within national tuberculosis programmes. The latter focuses on strengthening primary care provision in LMICs. The roll-out of these strategies in sub-Saharan Africa (sSA), where health systems remain fragmented and often dependent on externally funded vertical (communicable) disease control programmes, has been slow. Despite the longstanding existence of PAL, when patients with chronic respiratory symptoms present to health services in sSA, in practice they are faced with limited-service provision that often offers little in the way of alternative diagnostics and follow-on care, aside from an assessment for tuberculosis. While opportunities exist to build and integrate context-specific evidence-based healthcare interventions into tuberculosis services, it is essential to first understand the underlying cause/s of presumptive tuberculosis symptoms when they are not the result of active tuberculosis. The aim of this study was to undertake a systematic review of the evidence describing the number and nature of alternative final diagnoses among patients with presumed tuberculosis in sSA.

## Methods

### Search strategy and selection criteria

We performed a systematic review and meta-analysis of the evidence in accordance with PRISMA guidance. PROSPERO registration: CRD42018100004.^9^ We searched Ovid Medline, Embase, Cumulative Index of Nursing and Allied Health Literature (CINAHL) and the Cochrane library. The search strategy involved Medical Subject Heading and free text terms relating to the concepts of WHO tuberculosis symptoms (such as “chronic cough”, “fever” and “weight loss”), diagnostics (such as “diagnos*”, “sensitivity” and “specificity”), tuberculosis and utilised filters for North,^10^ East,^11^ South,^12^ West^13^ and Central Africa.^14^ The full Medline search strategy is provided in supplementary data and was modified for other databases. Human studies that met the inclusion criteria from inception to April 27, 2020 were included. No language restrictions were applied.

We included all studies (Diagnostic, Cohort and Observational) conducted in sSA enrolling adult (≥15 years old) patients with presumed tuberculosis presenting with symptoms (cough >2 weeks or any one of cough, fever, weight loss, night sweats or haemoptysis). Duplicate articles, research on non-human subjects, in-patient settings, articles reporting exclusively paediatric, extra-pulmonary, pregnant, prison or diabetic populations, and any studies irrelevant to tuberculosis and diagnostics not set in sSA were excluded. Narrative reviews, case reports, case series and studies reporting only smear microscopy diagnostics or screening with chest radiographs as opposed to symptoms were also excluded. This decision was based on recognition of poor access to high-quality radiography equipment and expert interpretation in sSA. Widespread use of low-quality radiography has been a barrier to large-scale programmatic use in tuberculosis programmes throughout sSA.

We screened citations of relevant articles and systematic reviews to identify additional studies. All articles identified by the initial search underwent title and abstract screening. Full-text review of potentially relevant articles was conducted. This was performed by two independent reviewers (SJ, FDD), where a third reviewer (CM) was called upon if a consensus could not be reached. If multiple studies used the same dataset or populations, we included the most comprehensive study with the largest number of participants and excluded the others. Multi-site studies were included where data from sSA sites were individually extractable from the total number of participants.

### Data analysis

Data extraction was performed by two independent reviewers (SJ and FDD) and compared, disagreements were resolved in the first instance by discussion and a third reviewer (CM) called upon if consensus could not be reached. A piloted standardised data extraction form was used to collect information from all eligible studies. All non-English language studies were translated using an online document translator.^15^

For each eligible study we extracted the year of publication, first authors name, mean or median age, proportion of male participants, study country, study setting (general or district hospital, local health centre or community), total number of participants eligible and included, diagnostic test used (culture or GeneXpert), number of patients with and without a diagnosis of tuberculosis disease (Bacteriologically confirmed or clinical) and their HIV rates, where available. Specific details of alternative diagnoses made, and their management were extracted. WHO Global Health Observatory data provided tuberculosis and HIV incidence estimates in-country during the years studies were undertaken and if they spanned more than a year the higher annual value used.

Included studies risk of bias was evaluated using a tool specifically for prevalence studies developed by the Joanna Briggs Institute.^16^ Each study was independently assessed according to ten items of methodological quality.

We used WHO case definitions for tuberculosis case reporting. These are bacteriologically confirmed tuberculosis cases and clinically diagnosed tuberculosis cases. All study participants included were tested for tuberculosis therefore clinically diagnosed tuberculosis cases in this review include patients with negative bacteriological results only and not patients that have not undergone testing.

All data analyses were done using R (version 4.0.2) and the metafor package version 2.4-0. We stratified random-effects meta-analyses of the proportion of patients with presumed tuberculosis found not to have tuberculosis by passive or active case finding, and whether cases found passively included clinically diagnosed cases. Meta-regression was used to assess the association between the proportion of patients with presumed tuberculosis subsequently found not to have tuberculosis and the proportion of clinically diagnosed tuberculosis cases, as well as with matched country-year estimates of per capita tuberculosis incidence and HIV prevalence.

## Results

Our search yielded 1799 articles. 246 duplicate articles were removed (Figure 1). After screening abstracts and titles, we excluded 1204 articles that were not relevant. After screening full texts, we excluded an additional 331 articles that did not meet the eligibility criteria. Therefore, 18 articles with 14527 participants from 10 African countries were included in this systematic review and meta-analysis. Bacteriologically confirmed tuberculosis refers to sputum culture positivity in all but one study^6^ that used Xpert MTB/RIF.

**Figure 1:**
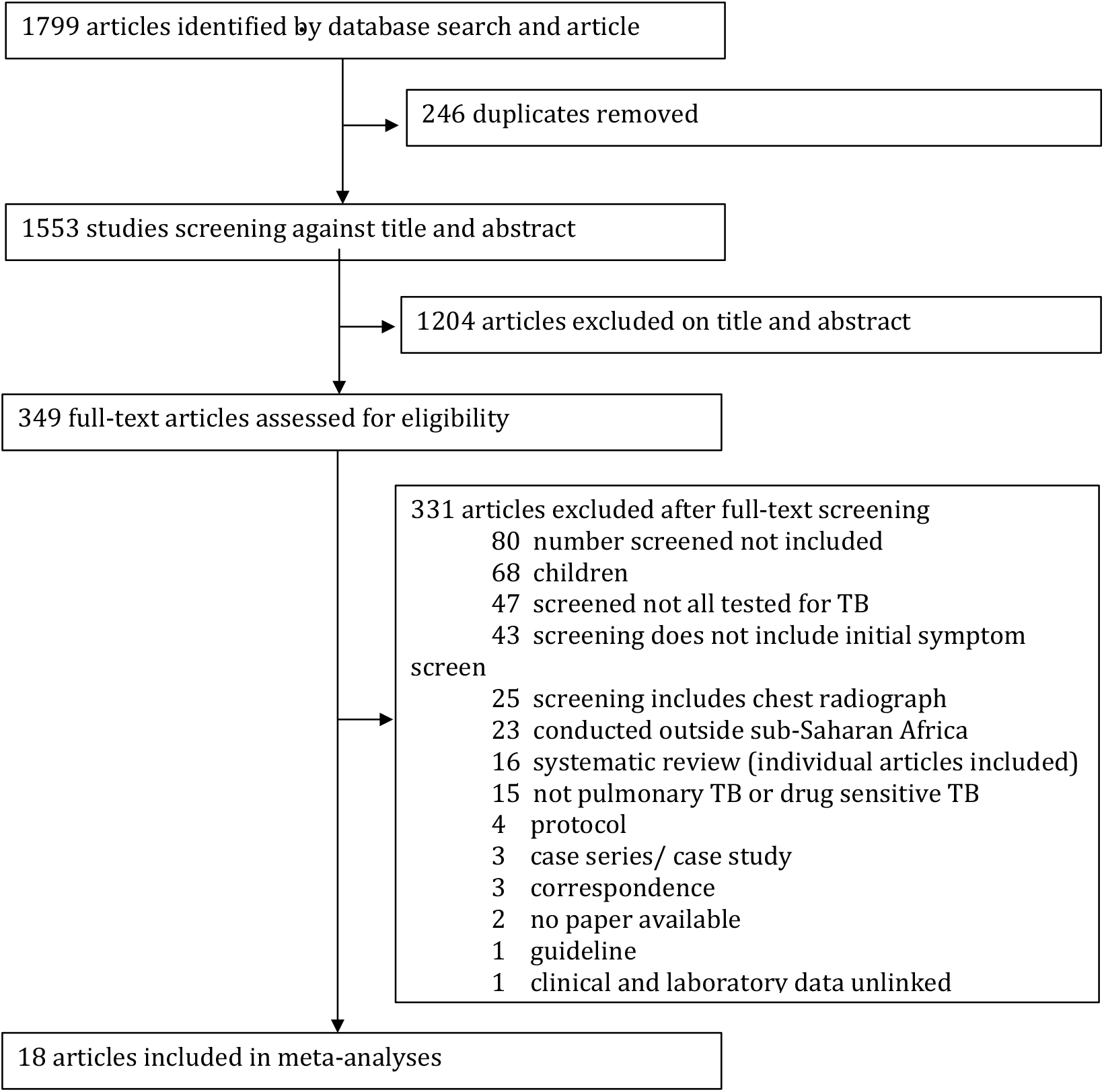
Study Selection

No studies were excluded following a risk of bias assessment (Supplementary data). All reporting 70% minimum study population coverage for tuberculosis diagnostic testing. Theron et al. (2011)^17^ and Ling et al. (2011)^18^ reported consecutive presumptive tuberculosis patient recruitment of 480 over 4yrs and 398 over 5yrs respectively. It was unclear how sampling was performed (breaks during sampling or sampled on certain days) and clinic sizes were not stated that could account for the long study periods with relatively low recruitment numbers.

There were seven studies including (Table 1a)^6,17,19-23^ and five studies not including (Table 1b)^18,24-27^ clinically diagnosed tuberculosis cases that used passive case finding strategies. Of the five studies (Table 1b) not including clinically diagnosed tuberculosis cases, only Dorman et al. (2018)^25^ did not document whether a clinical assessment was performed. Ling et al. (2011),^18^ Lawson et al. (2008),^27^ Hanrahan et al. (2014)^26^ and Cuevas et al. (2011)^24^ did perform a clinical assessment but reported no cases of clinically diagnosed tuberculosis. The proportion of patients with presumed tuberculosis subsequently found not to have tuberculosis increased with declining numbers of clinically diagnosed tuberculosis cases (P<0.0001).

**Table 1a.** Tuberculosis studies meeting inclusion criteria using passive case finding including clinically diagnosed tuberculosis cases

|  | Study Title | Study Type | Country | Age (median , IQR) | Male (%) | Setting | Presumptive TB (included/ eligible) | Diagnosed with Tuberculosis |  |  |  | Not Tuberculosis |  |
| --- | --- | --- | --- | --- | --- | --- | --- | --- | --- | --- | --- | --- | --- |
|  |  |  |  |  |  |  |  | Laboratory (bacteriological) | Clinical | Total | HIV | Total | HIV |
| Boehme et al. (2011)<sup>19</sup> | Feasibility, diagnostic accuracy, and effectiveness of decentralised use of the Xpert MTB/RIF test for diagnosis of tuberculosis and multidrug resistance: a multicentre implementation study | Cross-sectional Study | South Africa | 36, 29-46 | 51 | Health Centre | 1968/1968 | 473 | 824 | 1297 | NR | 671 | NR |
|  |  |  | Uganda | 32 26-38 | 54 | General Hospital | 307/307 | 146 | 17 | 163 | NR | 144 | NR |
| Bruchfeld et al. (2002)<sup>20</sup> | Evaluation of outpatients with suspected pulmonary tuberculosis in a high HIV prevalence setting in Ethiopia: Clinical, Diagnostic and epidemiological characteristics | Cross-sectional study | Ethiopia | 33<sup>‡</sup> | 56.3<sup>‡</sup> | General Hospital | 493/509 | 168 | 113 | 281 | 148 | 212 | 73 |
| Jayasooriya et al. (2019)<sup>6</sup> | The burden of non-TB lung disease presenting to TB clinics in the Gambia: Preliminary data in the Xpert MTB/rif era | Cross-sectional Study | The Gambia | 40<sup>‡</sup> 28-47 | 50<sup>‡</sup> | Research Clinic | 233/239 | 114 | 17 | 131 | 17 | 102 | 12 |
| Munyati et al. (2005)<sup>21</sup> | Chronic cough in primary health care attendees, Hasrare, Zimbabwe: diagnosis and impact of HIV infection | Cross-sectional Study | Zimbabwe | 33 | 48 | Health Centre | 544/550 | 184 | 50 | 234 | 207 | 310 | 247 |
| Nliwasa et al. (2016)<sup>22</sup> | The Sensitivity and Specificity of Loop-Mediated Isothermal Amplification (LAMP) Assay for Tuberculosis Diagnosis in Adults with Chronic Cough in Malawi | Cross-sectional Study | Malawi | 32<br>25-41 | 48 | Health Centre | 233/273 | 53 | 3 | 56 | 24 | 177 | 97 |
| Reither et al. (2010)<sup>23</sup> | Evaluation of diagnosis TB AG, a flow-through immunoassay for rapid detection of pulmonary tuberculosis | Cross-sectional Study | Tanzania | 36 | 47.4 | Research Clinic | 171/202 | 45 | 33 | 78 | 51 | 93 | 50 |
| Theron et al. (2011)<sup>17</sup> | Evaluation of the Xpert MTB/RIF assay for the diagnosis of pulmonary tuberculosis in a high prevalence setting | Cross-sectional Study | South Africa | 36<br>18-83<sup>†</sup> | 68 | Health Centre | 480/496 | 141 | 182 | 323 | 46 | 157 | 84 |

\* Age, mean (+/- SD), <sup>†</sup>(range) <sup>‡</sup> Not TB patients, <sup>§</sup>Not all tested, denominator, NR not recorded, HIV Human Immunodeficiency Virus

**Table 1b.** Tuberculosis studies meeting inclusion criteria using passive case finding not including clinically diagnosed tuberculosis cases

|  | Study Title | Study Type | Country | Age (median , IQR) | Male (%) | Setting | Presumptive TB (included/ eligible) | Diagnosed with Tuberculosis |  |  |  | Not Tuberculosis |  |
| --- | --- | --- | --- | --- | --- | --- | --- | --- | --- | --- | --- | --- | --- |
|  |  |  |  |  |  |  |  | Laboratory (bacteriological) | Clinical | Total | HIV | Total | HIV |
| Cuevas et al. (2011)<sup>24</sup> | A multi-country non-inferiority cluster randomized trials of frontloaded smear microscopy for the diagnosis of pulmonary tuberculosis | Cluster Randomised Trial | Ethiopia | 33.7* (+/-14.1) | 52.8 | Health Centre | 1770/1909 | 586 | 0 | 586 | 0 | 1184 | NR |
|  |  |  | Nigeria | 34.4* (+/-10.7) | 51.9 | Health Centre | 1196/1238 | 233 | 0 | 233 | 0 | 963 | NR |
| Dorman et al. (2018)<sup>25</sup> | Xpert MTB/RIF Ultra for detection of Mycobacterium tuberculosis and rifampicin resistance: a prospective multicentre diagnostic accuracy study | Cross-sectional Study | South Africa (Cape Town) | 41, 34-49 | 41 | District Hospital | 150/152 | 27 | NR | 27 | NR | 125 | NR |
|  |  |  | South Africa (Johannesburg) | 34, 30-43 | 63 | District Hospital | 234/234 | 74 | NR | 74 | NR | 160 | NR |
|  |  |  | Kenya | 33, 26-44 | 51 | District Hospital | 135/135 | 28 | NR | 28 | NR | 107 | NR |
|  |  |  | Uganda | 30, 26-39 | 64 | District Hospital | 181/181 | 67 | NR | 67 | NR | 114 | NR |
| Hanrahan et al. (2014)<sup>26</sup> | Xpert MTB/RIF as a measure of sputum bacillary burden. Variation by HIV status and immunosuppression | Cross-sectional Study | South Africa | 37, 29-46 | 38 | Health Centre | 2091/2406 | 406 | 0 | 406 | NR | 1685 | NR |
| Lawson et al. (2008)<sup>27</sup> | Clinical presentation of adults with pulmonary tuberculosis with and without HIV infection<br>in Nigeria | Cross-sectional Study | Nigeria | 33* (+/-10) | 61 | District Hospital | 1186/1321 | 731 | 0 | 731 | 329<sup>s</sup> /625 | 455 | 217<sup>s</sup> /377 |
| Ling et al<br>(2011)<sup>18</sup> | Are interferon-<br>gamma release<br>assays useful for<br>diagnosing active<br>tuberculosis in a<br>high-burden setting? | Cross-sectional<br>Study | South Africa | 40*<br>(+/-12) | 66 | Health<br>Centre | 395/500 | 138 | 0 | 138 | 43 | 257 | 65 |

\* Age, mean (+/- SD), †(range) ‡ Not TB patients, §Not all tested, denominator, NR not recorded, HIV Human Immunodeficiency Virus

There were four active case-finding studies without any clinically diagnosed tuberculosis cases (Table 2). Three studies were conducted in Ethiopia reporting clinical assessments, but no clinically diagnosed tuberculosis cases found.^28-31^ No clinical assessments were reported by Sekandi et al. in Uganda.^31^

**Table 2.**
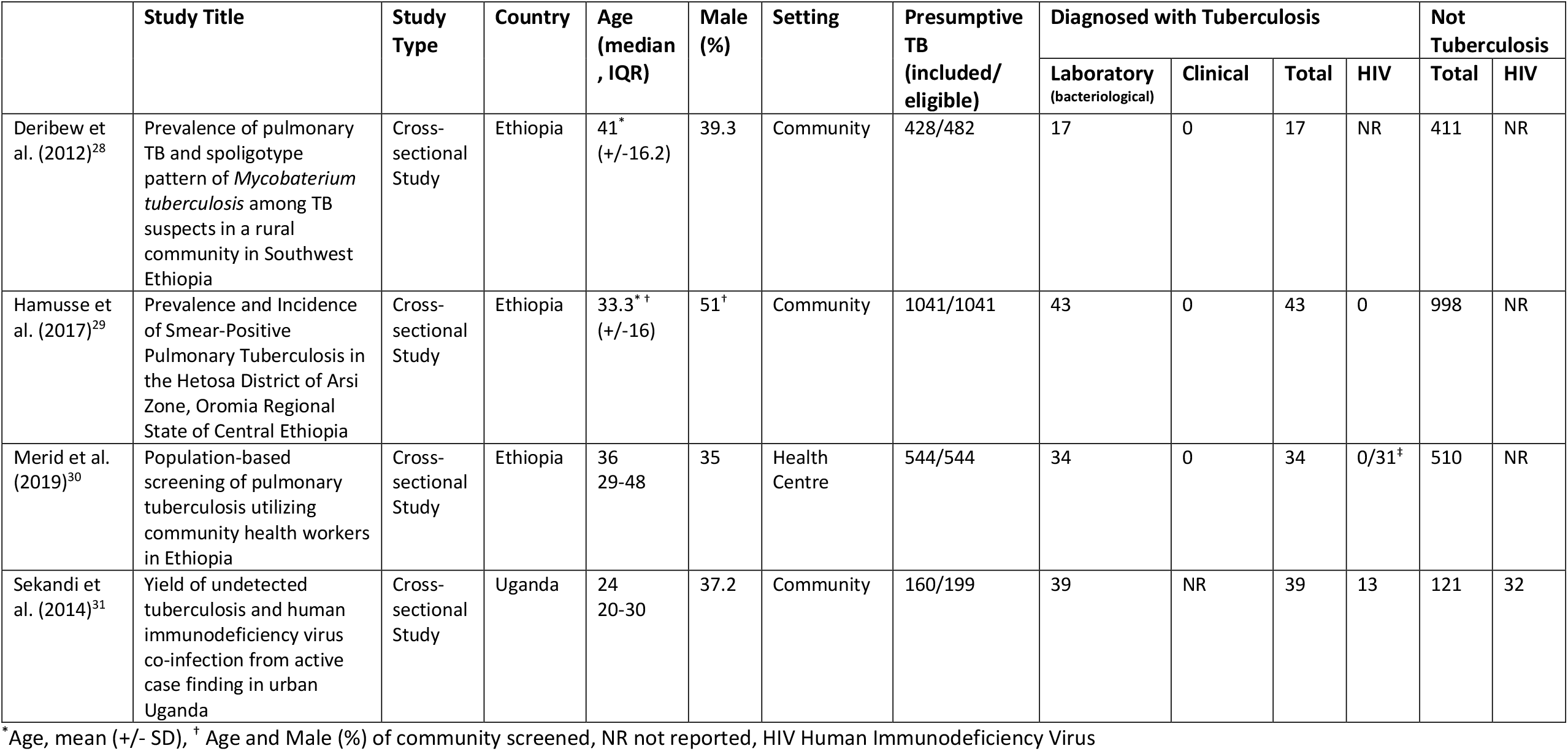
Tuberculosis studies meeting inclusion criteria using active finding not including clinically diagnosed tuberculosis cases

Figure 2 shows included studies and summary estimates grouped by passive and active case-finding. Passive case finding studies including clinically diagnosed tuberculosis cases (Table 1a) are shown in the top section of Figure 2 with estimates ordered by this proportion. The summary proportion of patients with presumed tuberculosis subsequently found not to have tuberculosis was lower in passive case finding studies that included clinically diagnosed tuberculosis cases (Table 1a) compared to those that did not (Table 1b), 48.5% (CI 38.4-56.7) versus 70.5% (CI 60.9-78.6) (Figure 2). Active case finding studies had higher proportions of patients with presumed tuberculosis subsequently found not to have tuberculosis, 92.7% (CI: 83.1-97.0) (Table 2, Figure 2). Among passive case finding studies, there were no statistically significant associations at the 0.05 level between country-year estimates of tuberculosis incidence or HIV prevalence. Heterogeneity was high (I^2>95% in all cases).

**Figure 2:**
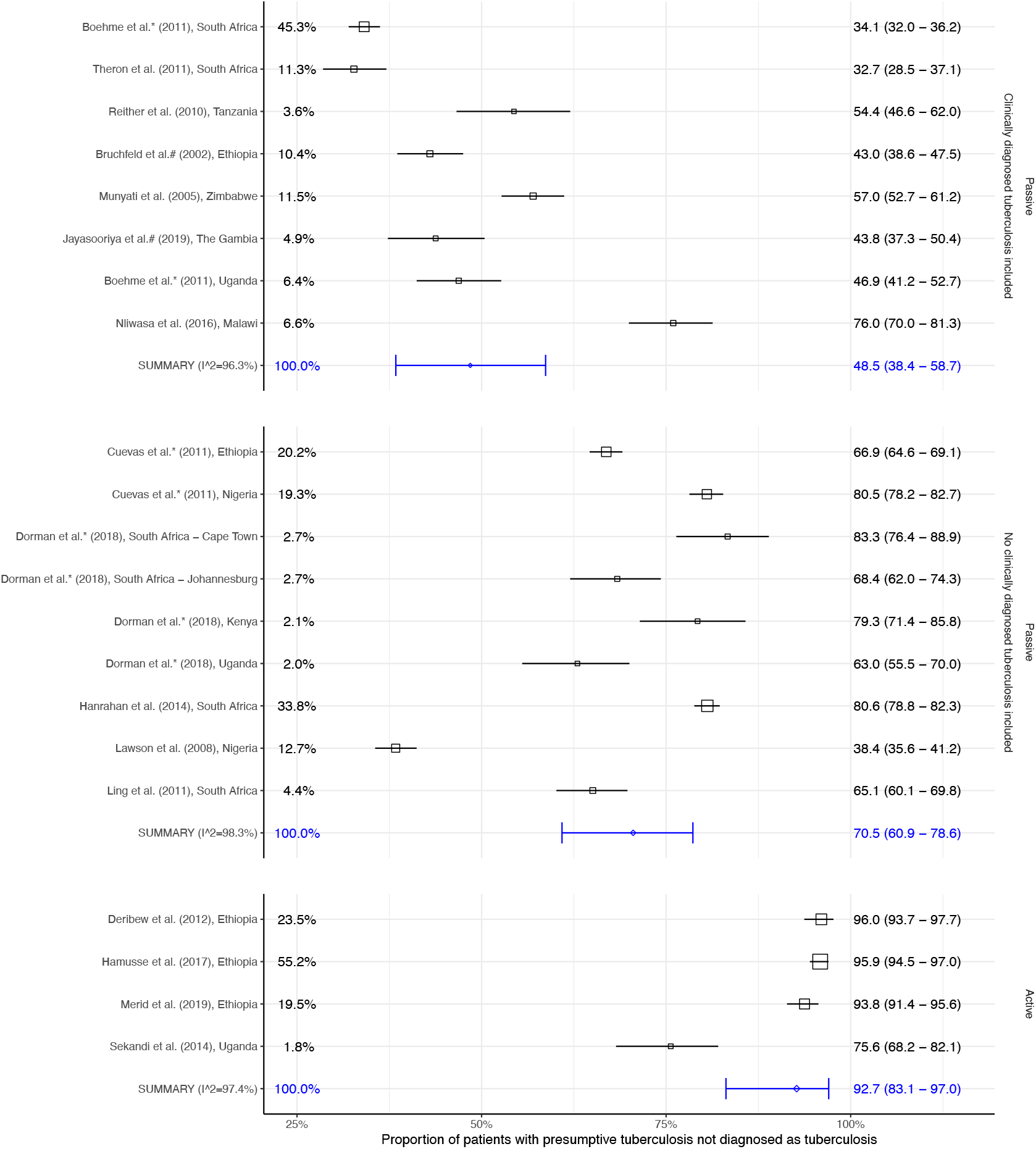
Random-effects meta-analyses of the proportion of patients with presumptive tuberculosis not diagnosed as tuberculosis. Studies are stratified by passive or active case finding. Passive case finding studies including clinically diagnosed tuberculosis are shown with estimates ordered by this proportion.

A further two articles included patients with presumed tuberculosis that were already smear negative on microscopy (Table 3). Affolabi et al. (2011)^32^ did not include and Huerga et al. (2012)^33^ included clinically diagnosed tuberculosis cases, with 89% and 61% of patients with presumed tuberculosis subsequently found not to have tuberculosis respectively.

**Table 3.** Tuberculosis studies of smear negative participants meeting inclusion criteria

|  | Study Title | Study Type | Country | Age (median , IQR) | Male (%) | Setting | Presumptive TB (included/ eligible) | Diagnosed with Tuberculosis |  |  |  | Not Tuberculosis |  |
| --- | --- | --- | --- | --- | --- | --- | --- | --- | --- | --- | --- | --- | --- |
|  |  |  |  |  |  |  |  | Laboratory (bacteriological) | Clinical | Total | HIV | Total | HIV |
| Affolabi et al. (2011)<sup>32</sup> | Smear-negative, culture-positive pulmonary tuberculosis among patients with chronic cough in Cotonou, Benin | Cross-sectional Study | Benin | NR | NR | General Hospital | 207/251 | 22 | 0 | 22 | 10 | 185 | 81 |
| Huerga et al. (2012)<sup>33</sup> | Performance of the 2007 WHO Alorithm to Diagnose Smear-Negative Pulmonary Tuberculosis in a HIV Prevalent Setting | Cross-sectional Study | Kenya | 34 (26-48) | 37.1 | District Hospital | 380/380 | 61 | 89 | 150 |  | 230 |  |

\*Age, mean (+/- SD), <sup>†</sup> Age and Male (%) of community screened, NR not reported, HIV Human Immunodeficiency Virus

Only five studies reported diagnoses other than active tuberculosis (Table 4).^6,20,21,26,33^ Two described non-tuberculous mycobacteria and one *Pneumocystis jirovecii* pneumonia as the only alternative diagnoses.^20,26,33^ Jayasooriya et al. (2019)^6^ and Munyati et al. (2005)^21^ described a range of diagnoses which were predominantly respiratory, but importantly non-respiratory diagnoses such as heart failure, malignancy and renal failure were noted. Neither study performed spirometry. Four out of the five studies reported management of patients with presumed tuberculosis subsequently found not to have tuberculosis, two stating as clinically indicated. Notably, Affolabi et al. (2011)^32^ and Huerga et al. (2012)^33^ reported giving empirical antibiotics to all patients subsequently found not to have active tuberculosis amounting to mass administration of antibiotics to 207 and 380 patients respectively. Only 10/18 (55%) studies recorded historical tuberculosis episodes, and none recorded the number of times individuals had undergone previous tuberculosis testing.

**Table 4.** Tuberculosis studies handling and reporting of patients with presumed tuberculosis found not to have tuberculosis

|  | Country | Diagnoses | Management | Past history of Tuberculosis | Previous Tuberculosis Testing | WHO estimated incidence (year of study) |  |
| --- | --- | --- | --- | --- | --- | --- | --- |
|  |  |  |  |  |  | Tuberculosis (per 100 000) | HIV (per 1000) |
| Affolabi et al. (2011)<sup>32</sup> | Benin | NR | 15days Erythromycin | NR | NR | 71 | 0.69 |
| Boehme et al. (2011)<sup>19</sup> | South Africa | NR | NR | NR | NR | 1260 | 10.29 |
|  | Uganda | NR | NR | NR | NR | 213 | 3.55 |
| Bruchfeld et al. (2002)<sup>20</sup> | Ethiopia | 8 Pneumocystis pneumonia | NR | 66* | NR | NR | 1.79 |
| Cuevas et al. (2011)<sup>24</sup> | Ethiopia | NR | NR | NR | NR | 296 | 0.44 |
|  | Nigeria | NR | NR | NR | NR | 219 | 0.79 |
| Deribew et al. (2012)<sup>28</sup> | Ethiopia | NR | NR | NR | NR | 282 | 0.41 |
| Dorman et al. (2018)<sup>25</sup> | South Africa (Cape Town) | NR | NR | 59 | NR | 805 | 5.45 |
|  | South Africa (Johanesburg) | NR | NR | 55 | NR | 805 | 5.45 |
|  | Kenya | NR | NR | 20 | NR | 348 | 1.17 |
|  | Uganda | NR | NR | 15 | NR | 201 | 1.89 |
| Hamusse et al. (2017)<sup>29</sup> | Ethiopia | NR | NR | NR | NR | 224 | 0.25 |
| Hanrahan et al. (2014)<sup>26</sup> | South Africa | 9 Non-tuberculous mycobacteria | NR | NR | NR | 1200 | 8.67 |
| Huerga et al (2012)<sup>33</sup> | Kenya | 11 Non-tuberculous mycobacteria | 5 days Amoxicillin | 92 | NR | 566 | 2.22 |
| Jayasooriya et al. (2019)<sup>6</sup> | The Gambia | 2 Malignancy:<br>2 Lung<br>1 Haematological | Clinically indicated | 16* | NR | 162 | 1.07 |
|  |  | 32 other respiratory tract infections |  |  |  |  |  |
|  |  | 8 Pneumonia |  |  |  |  |  |
|  |  | 4 Asthma |  |  |  |  |  |
|  |  | 2 Pleural effusions |  |  |  |  |  |
|  |  | 1 Lung abscess |  |  |  |  |  |
|  |  | 10 Heart failure |  |  |  |  |  |
|  |  | 2 Structural heart disease |  |  |  |  |  |
|  |  | 1 Ischaemic heart disease |  |  |  |  |  |
|  |  | 2 Chronic renal failure |  |  |  |  |  |
|  |  | 43 Unknown |  |  |  |  |  |
| Lawson et al. (2008)<sup>27</sup> | Nigeria | NR | NR | NR | NR | 219 | 0.91 |
| Ling et al (2011)<sup>18</sup> | South Africa | NR | NR | NR | NR | 1200 | 8.67 |
| Merid et al. (2019)<sup>30</sup> | Ethiopia | NR | NR | 151* | NR | 177 | 0.2 |
| Munyati et al. (2005)<sup>†21</sup> | Zimbabwe | 178 Other respiratory tract infections | Clinically indicated | 97 | NR | 607 | 8.67 |
|  |  | 87 bacterial Pneumonia |  |  |  |  |  |
|  |  | 34 Fibrotic lung disease:<br>28 Post-tuberculous disease<br>2 Idiopathic diffuse fibrosis |  |  |  |  |  |
|  |  | 26 Asthma |  |  |  |  |  |
|  |  | 8 Pneumocystis pneumonia |  |  |  |  |  |
|  |  | 5 Cryptococcosis |  |  |  |  |  |
|  |  | 15 Heart failure |  |  |  |  |  |
|  |  | 5 Malignancy:<br>3 Kaposi Sarcoma<br>1 primary bronchus<br>1 metastatic breast |  |  |  |  |  |
|  |  | 16 unknown |  |  |  |  |  |
| Nliwasa et al. (2016)<sup>22</sup> | Malawi | NR | NR | NR | NR | 261 | 3.2 |
| Reither et al (2010)<sup>23</sup> | Tanzania | NR | NR | NR | NR | 492 | 2.75 |
| Sekandi et al. (2014)<sup>31</sup> | Uganda | NR | NR | NR | NR | 217 | 3.7 |
| Theron et al. (2011)<sup>17</sup> | South Africa | NR | NR | 158 | NR | 1270 | 11.82 |

\* Past history of Tuberculosis in participants without Tuberculosis, † Participants diagnosed with multiple conditions, NR not reported

## Discussion

Our findings demonstrate that almost half of patients with presumed tuberculosis in sSA were not given a final diagnosis of active tuberculosis. While this proportion varied according to study it was not predicted by country incidence of tuberculosis or HIV. Only five of the identified studies attempted to characterise patients with presumed tuberculosis who were subsequently found not to have tuberculosis seeking alternative diagnoses.^6,20,21,26,33^ Of these, only two reported a breadth of alternative diagnoses.^6,21^ In both these cases, clinical judgement was used to decide on investigations performed, rather than a standardised approach, and in no instances was spirometry conducted. ^6,21^ Despite this cardiac pathology was described in both studies, consistent with a retrospective review of radiological findings in a Kenyan cohort identifying a wide variety of abnormalities unrelated to active tuberculosis.^34^

The proportion of patients with presumed tuberculosis subsequently found not to have tuberculosis was inversely associated with the number of clinically diagnosed tuberculosis cases. While this could imply overdiagnosis of active tuberculosis, through reliance on clinical judgement, it is important to note that LMICs have high rates of active tuberculosis.^4^ The lack of access to high quality health systems and diagnostics in sSA means there is likely to be a high burden of unrecognised diseases of all causes and unmet clinical need in the general population. ^35^ Therefore, patients with presumed tuberculosis – symptomatic by definition – risk having the true causes of their symptoms neglected if they are not due to active tuberculosis.^6,21^ An approach used in two included studies was mass administration of empirical antibiotics to several hundreds of patients with presumed tuberculosis subsequently not diagnosed with tuberculosis. With increasing antimicrobial resistance recognised as one of the biggest public health challenges of our time, nuanced strategies to mitigate against administering unnecessary antibiotics are vital. The implications for missing active tuberculosis are clear, yet those of incorrectly labelling people as having active tuberculosis, and/or missing other health conditions also need to be taken into consideration. Furthermore, how this relates to paediatric tuberculosis, where a high proportion of empirical tuberculosis treatment occurs, is unknown.

The few included studies that used active case finding strategies showed much higher proportions of patients with presumed tuberculosis subsequently found not to have tuberculosis than those that used passive case finding. This can in part be explained by active case finding studies reporting only bacteriologically confirmed tuberculosis cases. A WHO-commissioned systematic review reported general population community-based active case finding studies set in sSA^36^ detected bacteriologically (often smear) confirmed tuberculosis alone, and none reported clinically diagnosed tuberculosis cases. When comparing active with passive case finding studies that also reported only bacteriologically confirmed tuberculosis cases, the former still had a higher proportion of patients with presumed tuberculosis subsequently found not to have tuberculosis. These findings imply that active case finding strategies encounter more community members with unidentified health issues that have non-specific symptoms similar to those of active tuberculosis.

Systematic active screening of high-risk groups is a central component of the WHO End Tuberculosis Strategy and the aforementioned systematic review suggests that community-based active case-finding might be effective at detecting active tuberculosis early.^36^ However, the emphasis on active case-finding strategies in sSA should take into consideration patients with presumed tuberculosis subsequently found not to have tuberculosis. Improving the ability of local health systems to respond proactively to these patients, alongside clinically diagnosed tuberculosis cases is imperative.

Just over half of included studies captured prior histories of tuberculosis and none indicated how many times (if at all) patients had been previously tested for tuberculosis. Considering the former is important for assessing the risk of active tuberculosis in patients with presumed tuberculosis, recording this in future research is essential, as interpretation is limited by the incompleteness of these data. The impact of tuberculosis across the life course, evidenced by increased all-cause mortality post infection^37^ and post-tuberculosis lung disease, ^38^ in addition to implications for future test result interpretation, specifically Xpert MTB/RIF and Xpert MTB/RIF Ultra, also necessitate this. Moreover, patients with presumed tuberculosis subsequently found not to have tuberculosis will include some of the estimated 155 million patients globally alive today post tuberculosis^39^ and provide an opportunity to identify them allowing for provision of on-going care.

This work raises ethical issues around the inclusion of patients, not only those presumed to have tuberculosis, in research studies conducted in settings where limited primary care is available. This is especially true in view of rising chronic respiratory diseases, a major group of non-communicable diseases causing an estimated 3.9 million deaths in 2017 ^40^ and a disproportionately high burden in LMICs.^1^ It is critical that the duty of care afforded as a minimum to symptomatic unwell patients screening out of studies in such settings should be taken into consideration during research planning, offering for example in this case, enhanced standards of care and follow-up for patients subsequently found not to have tuberculosis. This will require improved collaboration between researchers and health system actors as well as greater consideration of the study participant’s health needs.

There are limitations to our review. We found only two studies with stated objectives to describe patients with presumed tuberculosis subsequently found not to have tuberculosis. Most studies were designed to capture patients with active tuberculosis and therefore understandably data on those not included as final study participants was not always comprehensive. In particular we highlight that where data was not recorded, it does not always equate to not being performed. However, this absence of data further supports our conclusion that there is a critical lack of reported data on patients with presumed tuberculosis subsequently found not to have tuberculosis.

This systematic review of the literature highlights that at least half of all patients with presumed tuberculosis attending services in sSA are not given a diagnosis of active tuberculosis; many not receiving any alternative diagnoses. In sSA 1.4million tuberculosis cases were notified in 2019, our data suggests this figure represents only half of all symptomatic patients presenting with presumptive tuberculosis.

Therefore, a further 1.4million may have accessed health systems but not received appropriate care. It is critical we address this by characterising the clinical needs among these hitherto neglected patients, in order to plan appropriate health system solutions. Future studies should explore patient experiences to better understand how these influence subsequent care seeking behaviours and health system engagement. Generating such data would help facilitate integration of services for non-communicable chronic respiratory diseases with tuberculosis programmes.

## Supporting information

Supplementary data - search strategy

Supplementary data - Risk of bias Table

## Data Availability

The protocol for this systematic review is available on PROSPERO: : CRD42018100004

## Contributors

SJ formulated the research questions with input from BK and KM

SJ, CB formulated the search strategy

SJ, FDD screened articles and data extracted with input from FC.

PJD synthesised data with input from SJ

All authors (SJ, FDD, CB, JB, BA, CM, BK, FC, PJD & KM) contributed to data interpretation and drafting the manuscript.

## Acknowledgements

We thank the NIHR Global Health Research Unit on Lung Health and tuberculosis in Africa at LSTM -“IMPALA” for helping to make this work possible. In relation to IMPALA (grant number 16/136/35) specifically: This research was funded by the National Institute for Health Research (NIHR) (IMPALA, grant reference 16/136/35) using UK aid from the UK Government to support global health research. The views expressed in this publication are those of the author(s) and not necessarily those of the NIHR or the UK Department of Health and Social Care.

PJD was supported by a fellowship from the UK Medical Research Council (MR/P022081/1); this UK funded award is part of the EDCTP2 programme supported by the European Union.

SJ was supported by an NIHR Clinical Lectureship Award.

