## Supplementary data - search strategy for "Patients with presumed tuberculosis in sub-Saharan Africa that are not diagnosed with tuberculosis: a systematic review and meta-analysis"

**Appendix 1: MEDLINE Search example**

Database: Ovid MEDLINE(R) and Epub Ahead of Print, In-Process & Other Non-Indexed Citations and Daily <1946 to April 27, 2020>

Search Strategy:

--------------------------------------------------------------------------------

1	(tubercul* adj3 symptom*).mp. [mp=title, abstract, original title, name of substance word, subject heading word, floating sub-heading word, keyword heading word, protocol supplementary concept word, rare disease supplementary concept word, unique identifier, synonyms] (889)

2	Cough/ (14630)

3	chronic cough.mp. [mp=title, abstract, original title, name of substance word, subject heading word, floating sub-heading word, keyword heading word, protocol supplementary concept word, rare disease supplementary concept word, unique identifier, synonyms] (3608)

4	exp Weight Loss/ or "weight loss".tw. (90993)

5	malaise.mp. [mp=title, abstract, original title, name of substance word, subject heading word, floating sub-heading word, keyword heading word, protocol supplementary concept word, rare disease supplementary concept word, unique identifier, synonyms] (6568)

6	fever/ (36688)

7	night sweats.mp. (1823)

8	Hemoptysis/ or (hemoptysis or haemoptysis).tw. (11303)

9	1 or 2 or 3 or 4 or 5 or 6 or 7 or 8 (160341)

10	exp "Sensitivity and Specificity"/ (533722)

11	sensitivity.tw. (709915)

12	specificity.tw. (416537)

13	((pre-test or pretest) adj probability).tw. (1941)

14	post-test probability.tw. (502)

15	predictive value$.tw. (95354)

16	likelihood ratio$.tw. (13524)

17	or/10-16 (1332010)

18	diagnos*.mp. (4528348)

19	active case.tw. (1030)

20	passive case.tw. (363)

21	sputum smear.tw. (2273)

22	sputum genexpert.tw. (4)

23	chest xray.tw. (54)

24	radiography,thoracic/ (30051)

25	screen*.tw. (650548)

26	diagnosis, differential/ (429446)

27	or/18-26 (4952361)

28	tuberculosis.mp. or TB.ti,ab. (250467)

29	17 or 27 (5777639)

30	exp africa, northern/ or (sudan* or western sahara* or algeria* or egypt* or libya* or morocc* or tunisia* or Cairo or Rabat or Casablanca or Tripoli or Algiers or Fes or Marrakesh or Tunis or Carthage or (Alexandria not (VA or Virginia)) or Tangier or Kairouan or Essaouira or Luxor or Bi zerte or "El Aaiun" or Sousse or Oran or Annaba or Constantine or Biskra or Chefchaoouen or Skikda or "Sharm El Sheikh" or Volubilis or "El Oued" or Meknes or Hippo Regius or Djemila or Sfax or Tataouine or port Said or "Ait Benhaddou" or Benghazi or Juba or Tamanrasette or merzouga or "El Djem" or oujda or Matmata or Ghat or Tabessa or Giza or Marj or Ifrane or "M'Hamid El Ghizlane" or Agadir or Tetouan or "Shubra El Kheima" or Tobruk or Khartoum or Nyala or Kassala or Ubayyid or Kosti or Wad Madani or Qadari f or Al - Fashir or Daein or Damazin or Geneina or Merowe or (north* adj2 africa*)).ti,ab. (67388)

31	exp Africa, Eastern/ or (((east* adj2 africa*) or British Indian Ocean Territory or Burundi* or Comoros or Djibouti* or Eritrea* or Ethiopia* or Kenya* or Madagascar or Malawi or Mauritius or Mayotte or Mozambique or Reunion or Rwanda* or Seychelles or Somalia* or Sudan* or Tanzania* or Uganda* or Zambia or Zimbabwe or Crozet Islands or Iles Crozet or Scattered Islands or Iles Eparses or Mwanza or Zanzibar or Eldoret or Morogoro or Hargeysa or Berbera or Nyeri or Mbeya or Machakos or Marka or Tabora or Iringa or Gondar or Meru or Geita or Musoma or Mtwara or Songea or Kigoma or Dese or Mek'ele or Bahir Dar or Jimma or Sinyanga or Korogwe or Nairobi or "Dar es Salaam" or Mombasa or Addis Ababa or Kampala or Kigali or Mogadishu or Dodomoa or Bujumbura or Nakuru or Anananarivo or Kisumu or Maputo or Asmara or Lusaka or Harare or Port Louis or Arusha or kitale or lilongwe or malindi or machakos or hargeisa or Bulawayo or Ruiru or Lamu or Kire Dawa or Kikuyu or naivasha or mwanza or tanga or nanyuki or voi or garissa or lodwar of kakamega or maralal or kitui or webuye or Axum or Nyahururu or Jinja or Kismayo or Namanga or Mumias or Moshi or Moroni or Lokichogio or Hola or Rwenzori Mountains or Lake Victoria or Puntland* or (Adiharush or Ali-Addeh or Alinjugur or Buramino or Dadaab or Dagahaley or Dollo Ado or Fugnido or Hagadera or Hilaweyn or Ifo or Kakuma or Kambioos or Kayaka II or Kobe or Kyangwali Nakivale or Nyarugusu or Wad Sherife or Bokolmanyo or Melkadida or Rwamanja)) adj5 (camp or refug*)).ti,ab. (54617)

32	exp africa, western/ or ((africa*adj2 west* or benin* or burkina fas* or cape verd* or cabo verd* or ivory coast or cote d'ivoire* or gambia* or ghana* or guinea* or bissau or liberia* or (mali not fowl) or malian or mauritania* or nigeria* or senegal* or sierra leon* or togo*).mp. or (Lagos or Accra or Abidjan or Dakar or Abobo or Abuja or Freetown or Ouagadougou or Conakry or Lome or Bamako or Cotonou or Kumasi or Monrovia or Ibadan or Kano or Port harcourt or Benin City or Porto Novo or Niamey or Yamoussoukro or Banjul or Timbuktu or Djenne or Abomeyu or Zaria or Tamale or Jos or Cape Coast or Maidugul or Aba or Gao or Calabar or Warri or Maiduguri or Bobo Dioulasso or Parakou or Djougou or Bohicon or Sekondi Takoradi or Sunyani or Obuasi or Teshie or Tema or Sikasso or Kalabankoro or Nouakchott or Dakhlet Nouadhibou or Benin City or Port Harcourt or Ilorin or Kaduna or Enugu or Ikorodu or Onitsha or Bauchi or Akure or Abeokuta or Sokoto or Bouake or Makeni or Kaduan or Sosgbo or Osogbo or Gombe or Ilesa or Badagry or makurdi or Sagamu or Iseyin or obbomosho or Awka or Ado Ekiti or Nsukka or Ikeja or Katsina or Okene or Lafia or Minna or Ondo city or Umuahia or Calabar or Yola or Pikine or Touba or Thies Nones or Saint Louis or Kolak or Ziguinch or (San Pedro not (Spain or Mexico or Argentina or California or United States or Italy)) or Bandama or Daloa or Owerri or Kandi or Ifi or Dakar or Ogbomosho or Divo or Korhogo)).ti,ab. (255692)

33	exp africa, central/ or ((africa adj2 central) or angola or cameroon* or chad.mp. or tchad.mp. or congo* or DRC or equatorial guinea* or gabon* or Sao Tome or Principe or Luanda or lobito or kuito or huambo or Malanje or Douala or Yaounde or Bamenda or Garoua of Bafoussam or Nganoundere or Maroua or Kouosseri or Buena or Kumba or N'Djamena or Moundou or Bangui or Bimbo or Brazzaville or Point Noire or Kinshasa or Lubumbashi or Leopoldville or Elizabethville or Mbuji Mayi or Bakwanga or Bukavu or Costermansville or Kananga or Luluabourg or Kisangani or Stanleyville or Tshikapa or Koalwezi or Likasi or Jadotville or Goma or Kikwit or Uvira or Bunia or Mbandaka or Coquilhatville or Matadi or Butembo or Kabinda or Mwene Ditu or Isiro or Paulis or Boma or Kindu or Bata or Malabo or Libreville).ti,ab. (31864)

34	exp africa, southern/ or ((africa* adj2 south*) or angola* or botswana* or lesotho* or malawi* or mozambiq* or namibia* or swaziland or zambia* or zimbabwe or Zulu or Tsonga or Xhosa or Swazi or Ndebele or Tswana or Sotho or Shona people or BaLunda or Mbundu or Ovimbundu or Chaga or Sukuma or Pretoria or Cape Town or Johannesburg or Durban or Port Elizabeth or Bloemfontein or Windhoek or Maseru or Pietermaritz or (Kimberley not Australia) or Nespruit or Soweto or Polokwane or Limpopo or Rustenburg or Mahikeng or Oudtshroom or Stellenbosch or Paarl or Gaborone or Luanda or Cabinda or Huambo or Lubango or Kuit or Malanje or Lobito or Lilongwe or Blantyre or Mzuzu or Maputo or Matola or Beira or Nampula or Chimoio or Nacala or Quelimane or Lusaka or Kitwe or Ndola or Kabwe or Copperbelt Harare or Bulawayo or Chitungwiza or Mutare or Masvingo or Monashonaland or Manicaland).ti,ab. (83002)

35	30 or 31 or 32 or 33 or 34 (470185)

36	9 and 28 and 29 and 35 (505)

37	9 and 27 and 28 and 35 (502)
