## Supplementary data - Risk of bias Table for "Patients with presumed tuberculosis in sub-Saharan Africa that are not diagnosed with tuberculosis: a systematic review and meta-analysis"

**Appendix 2**: JBI Risk of Bias Critical Appraisal Assessment Tool

|  | Was sample frame appropriate to address the target population? | Were study participants sampled in an appropriate way? | Was the sample size adequate? | Were the study subjects and the setting described in detail? | Was the data analysis conducted with sufficient coverage of the identified sample? | Were valid methods used for identification of the condition? | Was the condition measured in a standard, reliable way for all participants? | Was there appropriate statistical analysis? | Was the response rate adequate, and if not, was the low response rate managed appropriately? | Overall appraisal |
| --- | --- | --- | --- | --- | --- | --- | --- | --- | --- | --- |
| Affolabi et al. 2011* | 1 | 1 | 1 | 0 | 1 | 1 | 1 | 1 | 1 | INCLUDE |
| Boehme et al. 2011 | 1 | 0 | 0 | 1 | 1 | 1 | 1 | 1 | 1 | Uganda borderline sample size  Sampled set days  INCLUDE |
| Bruchfeld et al. 2002 | 1 | 0 | 1 | 1 | 1 | 1 | 1 | 1 | 1 | Convenience sampling  Sampled set days  INCLUDE |
| Cuevas et al. 2011 | 1 | 1 | 1 | 1 | 1 | 1 | 1 | 1 | 1 | INCLUDE |
| Deribew et al. 2012 | 1 | 1 | 1 | 1 | 1 | 1 | 1 | 1 | 1 | 11% not included  INCLUDE |
| Dorman et al. 2018 | 1 | 1 | 0 | 1 | 1 | 1 | 1 | 1 | 1 | INCLUDE |
| Hamusse et al. 2017 | 1 | 1 | 1 | 1 | 1 | 1 | 1 | 1 | 1 | INCLUDE |
| Hanrahan et al. 2014 | 1 | 1 | 1 | 1 | 1 | 1 | 1 | 1 | 1 | INCLUDE |
| Huerga et al. 2012* | 1 | 1 | 1 | 1 | 1 | 1 | 1 | 1 | 1 | INCLUDE |
| Jayasooriya et al. 2019 | 1 | 1 | 0 | 1 | 1 | 1 | 1 | 1 | 1 | INCLUDE |
| Lawson et al. 2008 | 1 | 1 | 1 | 1 | 1 | 1 | 1 | 1 | 1 | 11% not included  INCLUDE |
| Ling et al. 2011 | 1 | 1 | 1 | 1 | 0 | 1 | 1 | 1 | 1 | Sampled over 3years  21% not included  INCLUDE |
| Merid et al. 2019 | 1 | 1 | 1 | 1 | 1 | 1 | 1 | 1 | 1 | INCLUDE |
| Munyati et al. 2005 | 1 | 0 | 1 | 1 | 1 | 1 | 1 | 1 | 1 | Convenience sampling  INCLUDE |
| Nliwasa et al. 2016 | 1 | 0 | 1 | 1 | 1 | 1 | 1 | 1 | 1 | Borderline sample size  INCLUDE |
| Reither et al. | 1 | 2 | 0 | 0 | 1 | 1 | 1 | 1 | 1 | Limited setting detail  INCLUDE |
| Sekandi et al. 2014 | 1 | 0 | 1 | 1 | 1 | 1 | 1 | 1 | 1 | Small sample size  INCLUDE |
| Theron et al. 2011 | 0 | 2 | 1 | 1 | 1 | 1 | 1 | 1 | 2 | Sampled over 3years  INCLUDE |

0=No, 1=Yes, 2=Unclear 3=Not Applicable ^*^ Smear negative studies
